# Ductal Carcinoma In Situ with Diffuse Growth Distribution: A Potentially Lethal Subtype of “Preinvasive” Disease

**DOI:** 10.1101/2020.09.17.20196931

**Authors:** Thomas J O’Keefe, Olivier Harismendy, Anne M Wallace

**Affiliations:** Division of Breast Surgery And The Comprehensive Breast Health Center, University of California San Diego Mailbox 0819, 3855 Health Sciences Dr, La Jolla, CA 92037; Moores Cancer Center and Division of Biomedical Informatics, Department of Medicine, University of California San Diego 3855 Health Sciences Dr, La Jolla, CA 92037

## Abstract

**PURPOSE:** The current trend in ductal carcinoma in situ (DCIS) research is towards treatment de-escalation. Problematically, prognostic indicators for patients at high risk of breast cancer mortality (BCM), rather than recurrence, have not been identified. We aim to identify prognostic factors for the development of metastatic disease and mortality.

**EXPERIMENTAL DESIGN:** Patients diagnosed with DCIS in a local cancer registry as well as in the National Cancer Database (NCDB) and the Surveillance, Epidemiology and End Results (SEER) program were assessed for factors prognostic of metastatic disease, overall, and breast-cancer specific survival. Cox and competing risks regressions were developed.

**RESULTS:** Among 5 patients who developed distant metastatic disease in the cancer registry, 3 had identifiable growth distribution; all 3 were diffuse type. None had in-breast invasive or DCIS recurrences before metastasis. In NCDB and SEER, cumulative incidence of any cause mortality (ACM) and BCM at 10 years was 12%/5.0% for diffuse lesions; 8%/3.6% for patients with microinvasive disease, 7.4%/2.3% for lesions >5 cm, 5.6%/1.4% for lesions 2-5 cm and 5.5%/1.5% for lesions <2 cm. Multivariate hazard ratios for ACM in NCDB and BCM in SEER were 2.0 and 5.3 (p=0.03 and 0.02, respectively). Among patients with diffuse lesions, cumulative incidence ACM at 10 years was 15.0% among those undergoing unilateral mastectomy vs. 2.5% among those undergoing bilateral mastectomy (p=0.11).

**CONCLUSION:** Diffuse DCIS represents an uncommon but deadly subtype for whom treatment escalation, rather than de-escalation, is likely necessary. Further studies elucidating the mechanism of metastasis and best treatment course are needed.

## Introduction

### Evolution of DCIS Management and the “Mastectomy Blind Spot”

Ductal carcinoma in situ (DCIS) is a commonly diagnosed neoplastic process of the breast that encompasses a heterogeneous group of lesions which vary in histopathologic features and outcomes [1]. Prior to the 1980s, mastectomy was standard of care for DCIS because of the potential of DCIS to progress to invasive disease [2, 3]. After randomized trials demonstrated that mastectomy did not result in a difference in outcomes for invasive breast cancer patients relative to breast conservation therapy, lumpectomy was adopted for DCIS [2, 3].

Treatment was then further refined in response to multiple DCIS randomized trials which demonstrated that the addition of radiation therapy (RT) to lumpectomy reduced ipsilateral DCIS and invasive recurrences, and among estrogen receptor (ER) positive patients endocrine therapy (ET) such as tamoxifen reduced some combination of in situ and invasive recurrences in either breast [2, 3]. Notably neither of these adjuvant therapies resulted in a reduction in breast cancer mortality (BCM) [2, 3]. Importantly, these trials investigated patients undergoing lumpectomy, not mastectomy, in spite of the fact that mastectomies represent between 27-56% of the surgeries received by DCIS patients [4-10].

### Current DCIS Research Trends

As a result of the low BCM rate observed among DCIS patients, as well as the fact that prevention of recurrences has not been shown to reduce BCM [11], both treatment and research currently emphasize de-escalation of treatment and identification of low risk patients, with the most common outcome of interest being invasive recurrence [3, 12-14]. Efforts to identify DCIS patients at high risk, and in particular at high risk of BCM, have been less robust, despite the longstanding existence of indirect evidence that some DCIS may have the potential to metastasize [11, 15-22]. It remains unclear whether metastases after diagnosis of DCIS are secondary to occult disease, temporally or spatially missed foci of invasion, or a different mechanism altogether such as intraductal angiogenic processes.

### DCIS Growth Distribution

DCIS lesion size is a known independent predictor of residual disease [23, 24]. Multifocality, when multiple foci of disease arise within the same quadrant of the breast, has also been linked with prognosis among patients with DCIS relative to unifocal disease, in which there is a single focus of DCIS [25]. We will use the term multifocal to refer to both multifocal disease and multicentric disease, the latter of which refers to multiple foci of disease arising in different quadrants of the breast. Bonnier et al. found in a retrospective study that among DCIS patients undergoing lumpectomy with RT, multifocal disease was significantly associated with decreased recurrence free and metastatic free survival relative to patients with unifocal disease, though this association was not identified among patients undergoing mastectomy [26]. Swedish pathologist Tibor Tot identified a third spatial configuration of DCIS beyond unifocal and multifocal disease which he dubbed diffuse disease, referring to tumors spread out over a large area with no distinct tumor edge [27]. He later demonstrated that this diffuse distribution as well as multifocal disease were associated with BCM relative to unifocal lesions in an analysis including mostly mixed invasive and in situ but some pure in situ lesions [27-29].

### Aims

We sought to investigate factors that affect development of metastatic disease, breast cancer mortality, and overall mortality in patients diagnosed with DCIS at our institution, in SEER, and NCDB, with emphasis on the impact of growth distribution (GD).

## Methods

This study was conducted in accordance with U.S. Common Rule. The UCSD HRPP/IRB approved the UCSD registry portion of the study and deferred need for approval of the SEER and NCDB portions of the study due to use of public, de-identified data. Informed consent was not obtained for the cancer registry patients due to the large number of patients involved and was not performed for the SEER and NCDB patients due to de-identified data.

### Data Extraction

UCSD Cancer Registry data was collected for all breast cases from January 1, 2000 to March 21, 2019. Patients were excluded if they had invasive disease, were male, had only lumpectomy for their DCIS, had a prior cancer event, or had missing pathology reports (Figure 1).

**Figure 1.**
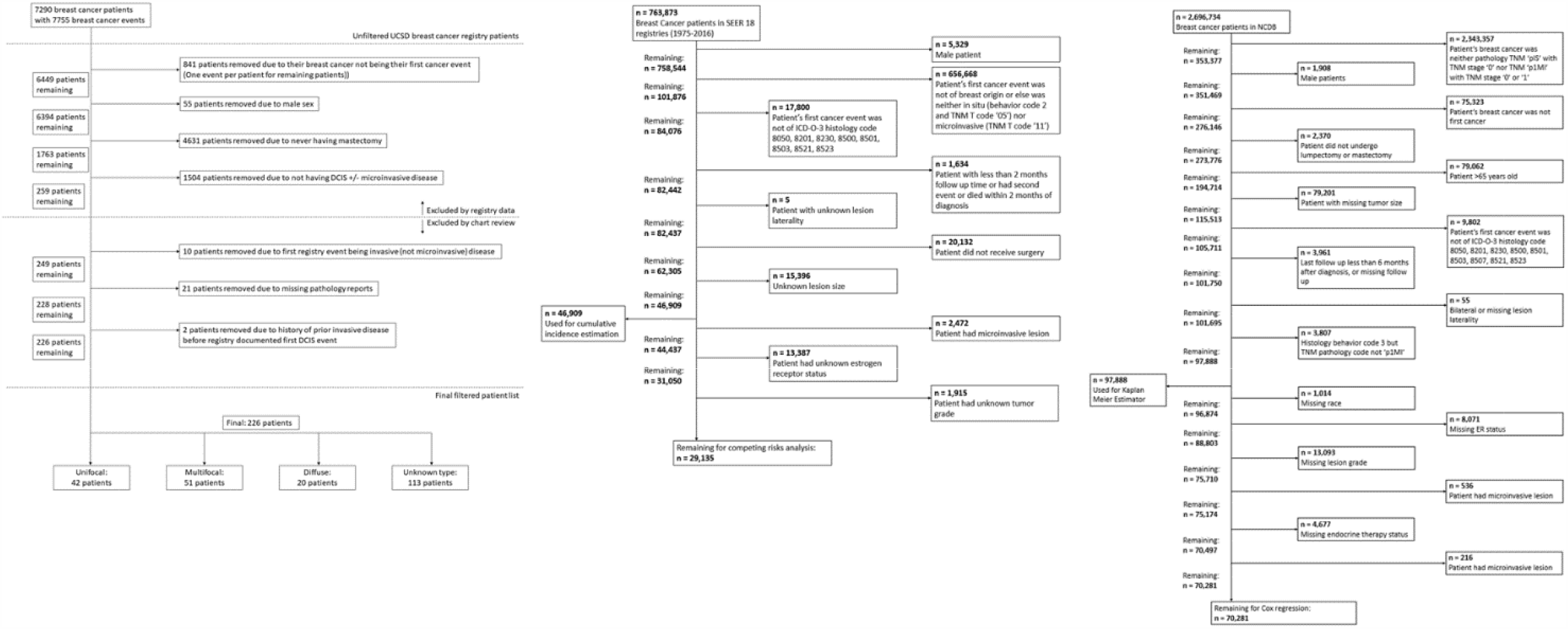
Sequential application of exclusion criteria to patients in UCSD cancer registry.

SEER 18 data was used from cases diagnosed in 1975-2016, and patients were excluded if they were male, had a prior cancer event, had non-DCIS histology ICD-O-3 code, had less than 2 months follow-up, had bilateral or missing lesion laterality, did not undergo surgery, or had missing lesion size, grade, or ER status (Figure 1).

NCDB data was filtered in a similar manner to SEER data, except we also excluded patients >65 and patients with missing endocrine therapy status, and we used a follow up time cut off of 6 months instead of 2 (Figure 1).

For SEER and NCDB, patients with ER positive and borderline disease were categorized together, and patients of non-white, non-black race with non-missing race were categorized together.

### Growth Distribution Classification

For UCSD patients, the growth distribution of the lesions were classified into the following groups: unifocal, multifocal, and diffuse. In cases in which the pathology report stated that the lesion was unifocal, it was classified as such. In cases in which the lesion had descriptive characteristics of numerous, scattered, or a multitude of small or microscopic foci, with or without an associated larger focal lesion or lesions, the lesion was classified as diffuse. In cases in which the previously mentioned descriptive terms were not applied and the lesion was classified on the pathology report as multifocal or multicentric, the lesion was classified as multifocal. Largest contiguous size of disease was documented for all lesion types and extent of disease was recorded for multifocal and diffuse type lesions.

For SEER and NCDB patients, lesion growth distribution is not generally documented, but a tumor size code exists for diffuse type disease (‘998’ in both). For both of these databases, lesion size was categorized as follows: ≤2 cm, 2 – 5 cm, >5 cm, and diffuse. For cumulative incidence estimators including microinvasive disease, patients with microinvasive disease of any size were grouped into a fifth category.

### Statistical Analysis

Patients of different growth distributions at UCSD were compared using Chi-Square and t-test. Kaplan-Meier method was used to estimate cumulative incidence for development of metastatic disease among UCSD patients and for overall survival (OS) among NCDB patients and log-rank test was used to compare groups. Cox regression with and without time-varying coefficients in the form of step functions were used to approximate hazard ratios [HR] for these respective data and outcomes.

For SEER, competing risks cumulative incidence was approximated with the competing events including BCM and non-breast mortality. An additional analysis was performed in which BCM in the absence and presence of an in breast recurrence were treated as distinct competing risks. Competing risks regression by Fine and Gray method [30], again with and without time-varying coefficients in the form of step functions, was performed to approximate HR for BCM.

We tested the proportional hazards assumption both graphically and numerically for both Cox and competing risks regressions, with addition of time-varying coefficients for covariates violating the assumption. P values were calculated as two-sided and statistical significance was declared for p less than 0.05. All statistical analysis was performed in R (version 3.5.1, R Foundation for Statistical Computing, Vienna, Austria) using RStudio (Version 1.1.463) and packages “tidyverse” (Version 1.3.0), “survival” (Version 3.1-7), “survminer” (Version 0.4.6), and “cmprsk” (Version 2.2-9).

## Results

### UCSD Cancer Registry

A total of 226 patients met the inclusion criteria, of whom 113 could be confidently classified according to GD. 42 (37.2%) had unifocal, 51 (45.1%) had multifocal, and 20 (17.7%) had diffuse lesions (Table 1). Median follow up time was 7.1 years (interquartile range 4.1-11.7 years). Median age at diagnosis was 49.

**Table 1.**
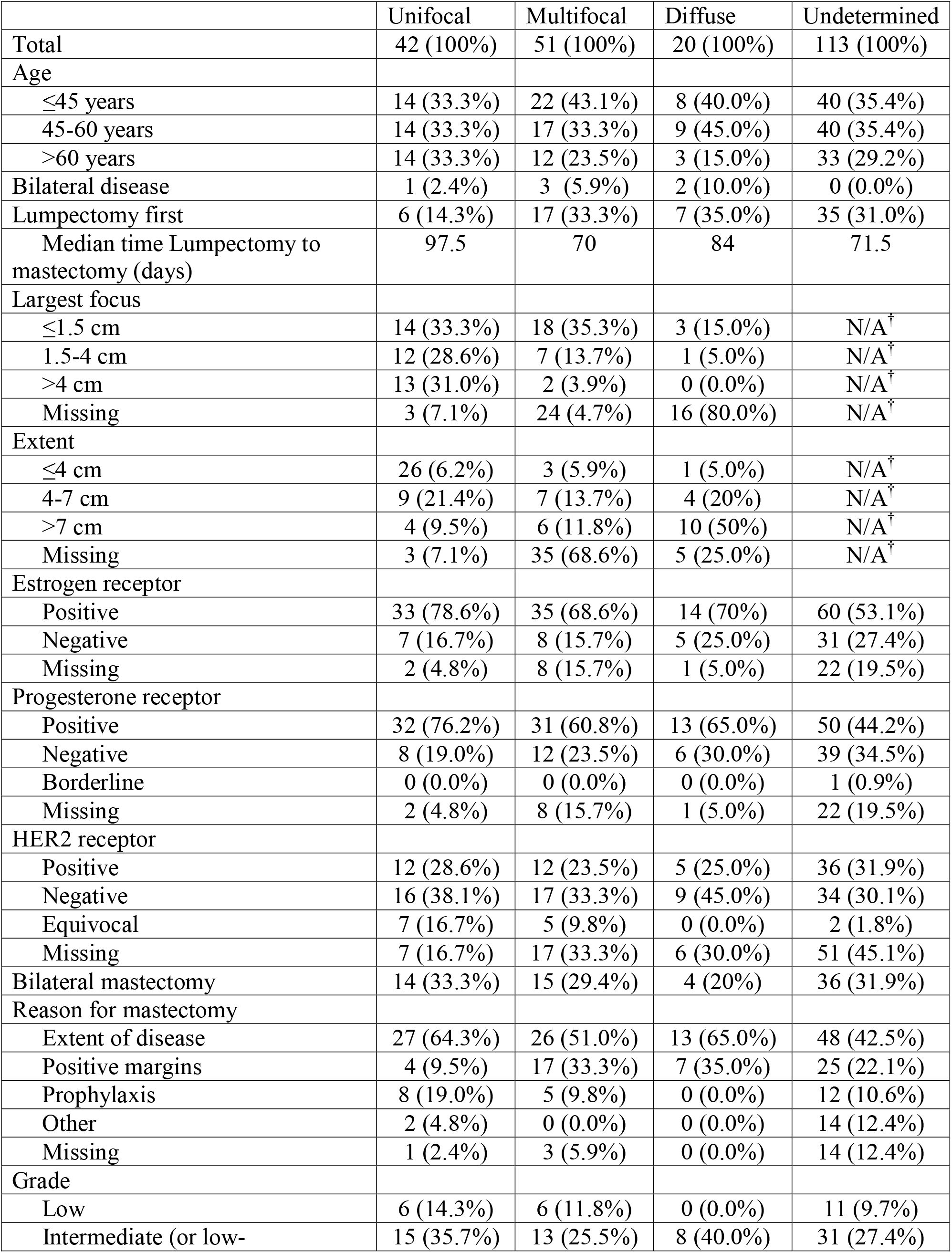
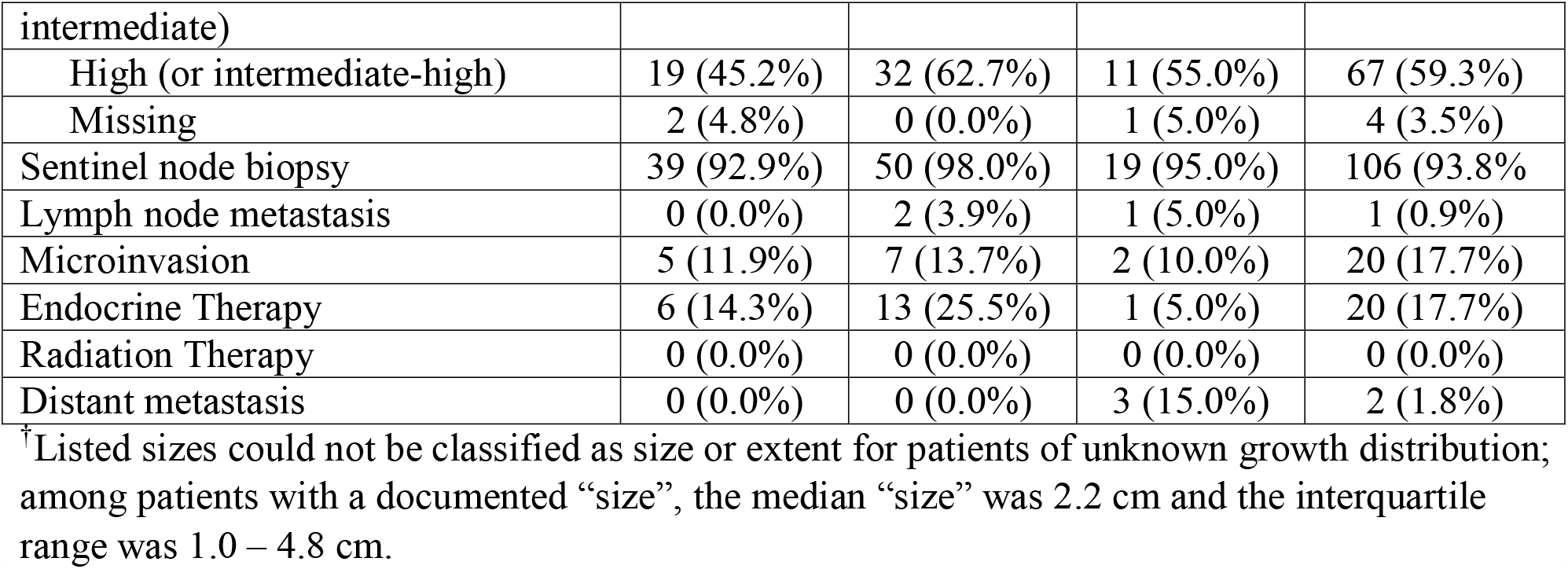
UCSD cancer registry characterization of DCIS patients grouped by growth distribution.

|  | Unifocal | Multifocal | Diffuse | Undetermined |
| --- | --- | --- | --- | --- |
| Total | 42 (100%) | 51 (100%) | 20 (100%) | 113 (100%) |
| Age |  |  |  |  |
| ≤45 years | 14 (33.3%) | 22 (43.1%) | 8 (40.0%) | 40 (35.4%) |
| 45-60 years | 14 (33.3%) | 17 (33.3%) | 9 (45.0%) | 40 (35.4%) |
| >60 years | 14 (33.3%) | 12 (23.5%) | 3 (15.0%) | 33 (29.2%) |
| Bilateral disease | 1 (2.4%) | 3 (5.9%) | 2 (10.0%) | 0 (0.0%) |
| Lumpectomy first | 6 (14.3%) | 17 (33.3%) | 7 (35.0%) | 35 (31.0%) |
| Median time Lumpectomy to mastectomy (days) | 97.5 | 70 | 84 | 71.5 |
| Largest focus |  |  |  |  |
| ≤1.5 cm | 14 (33.3%) | 18 (35.3%) | 3 (15.0%) | N/A<sup>†</sup> |
| 1.5-4 cm | 12 (28.6%) | 7 (13.7%) | 1 (5.0%) | N/A<sup>†</sup> |
| >4 cm | 13 (31.0%) | 2 (3.9%) | 0 (0.0%) | N/A<sup>†</sup> |
| Missing | 3 (7.1%) | 24 (4.7%) | 16 (80.0%) | N/A<sup>†</sup> |
| Extent |  |  |  |  |
| ≤4 cm | 26 (6.2%) | 3 (5.9%) | 1 (5.0%) | N/A<sup>†</sup> |
| 4-7 cm | 9 (21.4%) | 7 (13.7%) | 4 (20%) | N/A<sup>†</sup> |
| >7 cm | 4 (9.5%) | 6 (11.8%) | 10 (50%) | N/A<sup>†</sup> |
| Missing | 3 (7.1%) | 35 (68.6%) | 5 (25.0%) | N/A<sup>†</sup> |
| Estrogen receptor |  |  |  |  |
| Positive | 33 (78.6%) | 35 (68.6%) | 14 (70%) | 60 (53.1%) |
| Negative | 7 (16.7%) | 8 (15.7%) | 5 (25.0%) | 31 (27.4%) |
| Missing | 2 (4.8%) | 8 (15.7%) | 1 (5.0%) | 22 (19.5%) |
| Progesterone receptor |  |  |  |  |
| Positive | 32 (76.2%) | 31 (60.8%) | 13 (65.0%) | 50 (44.2%) |
| Negative | 8 (19.0%) | 12 (23.5%) | 6 (30.0%) | 39 (34.5%) |
| Borderline | 0 (0.0%) | 0 (0.0%) | 0 (0.0%) | 1 (0.9%) |
| Missing | 2 (4.8%) | 8 (15.7%) | 1 (5.0%) | 22 (19.5%) |
| HER2 receptor |  |  |  |  |
| Positive | 12 (28.6%) | 12 (23.5%) | 5 (25.0%) | 36 (31.9%) |
| Negative | 16 (38.1%) | 17 (33.3%) | 9 (45.0%) | 34 (30.1%) |
| Equivocal | 7 (16.7%) | 5 (9.8%) | 0 (0.0%) | 2 (1.8%) |
| Missing | 7 (16.7%) | 17 (33.3%) | 6 (30.0%) | 51 (45.1%) |
| Bilateral mastectomy | 14 (33.3%) | 15 (29.4%) | 4 (20%) | 36 (31.9%) |
| Reason for mastectomy |  |  |  |  |
| Extent of disease | 27 (64.3%) | 26 (51.0%) | 13 (65.0%) | 48 (42.5%) |
| Positive margins | 4 (9.5%) | 17 (33.3%) | 7 (35.0%) | 25 (22.1%) |
| Prophylaxis | 8 (19.0%) | 5 (9.8%) | 0 (0.0%) | 12 (10.6%) |
| Other | 2 (4.8%) | 0 (0.0%) | 0 (0.0%) | 14 (12.4%) |
| Missing | 1 (2.4%) | 3 (5.9%) | 0 (0.0%) | 14 (12.4%) |
| Grade |  |  |  |  |
| Low | 6 (14.3%) | 6 (11.8%) | 0 (0.0%) | 11 (9.7%) |
| Intermediate (or low- | 15 (35.7%) | 13 (25.5%) | 8 (40.0%) | 31 (27.4%) |
| intermediate) |  |  |  |  |
| High (or intermediate-high) | 19 (45.2%) | 32 (62.7%) | 11 (55.0%) | 67 (59.3%) |
| Missing | 2 (4.8%) | 0 (0.0%) | 1 (5.0%) | 4 (3.5%) |
| Sentinel node biopsy | 39 (92.9%) | 50 (98.0%) | 19 (95.0%) | 106 (93.8%) |
| Lymph node metastasis | 0 (0.0%) | 2 (3.9%) | 1 (5.0%) | 1 (0.9%) |
| Microinvasion | 5 (11.9%) | 7 (13.7%) | 2 (10.0%) | 20 (17.7%) |
| Endocrine Therapy | 6 (14.3%) | 13 (25.5%) | 1 (5.0%) | 20 (17.7%) |
| Radiation Therapy | 0 (0.0%) | 0 (0.0%) | 0 (0.0%) | 0 (0.0%) |
| Distant metastasis | 0 (0.0%) | 0 (0.0%) | 3 (15.0%) | 2 (1.8%) |

<sup>†</sup>Listed sizes could not be classified as size or extent for patients of unknown growth distribution; among patients with a documented “size”, the median “size” was 2.2 cm and the interquartile range was 1.0 – 4.8 cm.

Subsequent distant metastatic disease (DMD) or sentinel lymph node involvement at time of DCIS operation (SLNI) were identified in 9 (4.0%) of the 226 patients (Table 2), including 5 (2.2%) patients who developed DMD and 4 (1.8%) who had SLNI. Median age at diagnosis for the SLNI/DMD cases was lower than the overall group at 42 years. All 3 DMD patients classified by GD were had diffuse GD. No DMD patients had an intervening in breast invasive nor in situ recurrence, nor positive margins, and none of the 4 undergoing genetic testing had a positive result. Furthermore, none of the patients who developed DMD had involvement of their SLNB at the time of surgery, and none of the patients who had SLNI at the time of surgery went on to develop metastatic disease.

**Table 2.**
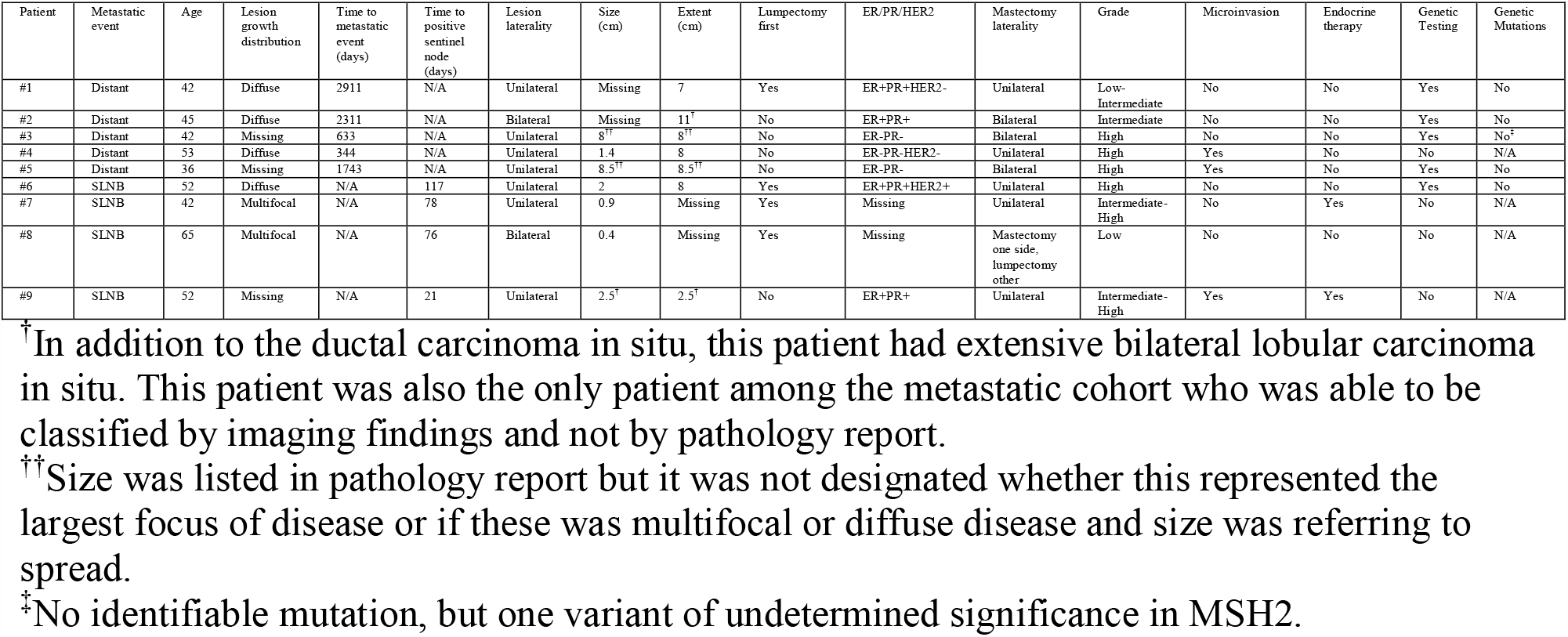
Characterization of patients within the cohort who developed subsequent distant metastatic disease or had local lymph node involvement at the time of surgery.

Given the close association of diffuse GD with development of DMD in this registry study, but the very small number of patients involved preventing meaningful survival analysis, we turned to national cancer databases, NCDB and SEER, to further evaluate the effects of diffuse type disease.

### National Cancer Database

A total of 70,281 patients met inclusion criteria for the final Cox regression. 52,859 patients had lesion sizes ≤ 2 cm, 13,097 had lesions 2-5 cm, 4,137 patients had lesions > 5 cm, and 188 patients had lesion sizes classified as diffuse. Median follow up time was 4.3 years (interquartile range 2.4-6.6 years). Median age at diagnosis was 53.

Cumulative incidence of any cause mortality (ACM) at 10 years was 12% for patients with diffuse lesions, 7.4% for lesions >5 cm, 5.6% for lesions 2-5 cm and 5.5% for lesions ≤ 2 cm (Figure 2). When considered as a separate group, patients with microinvasive disease, regardless of tumor size, had a 10 year cumulative incidence of ACM of 8%.

**Figure 2.**
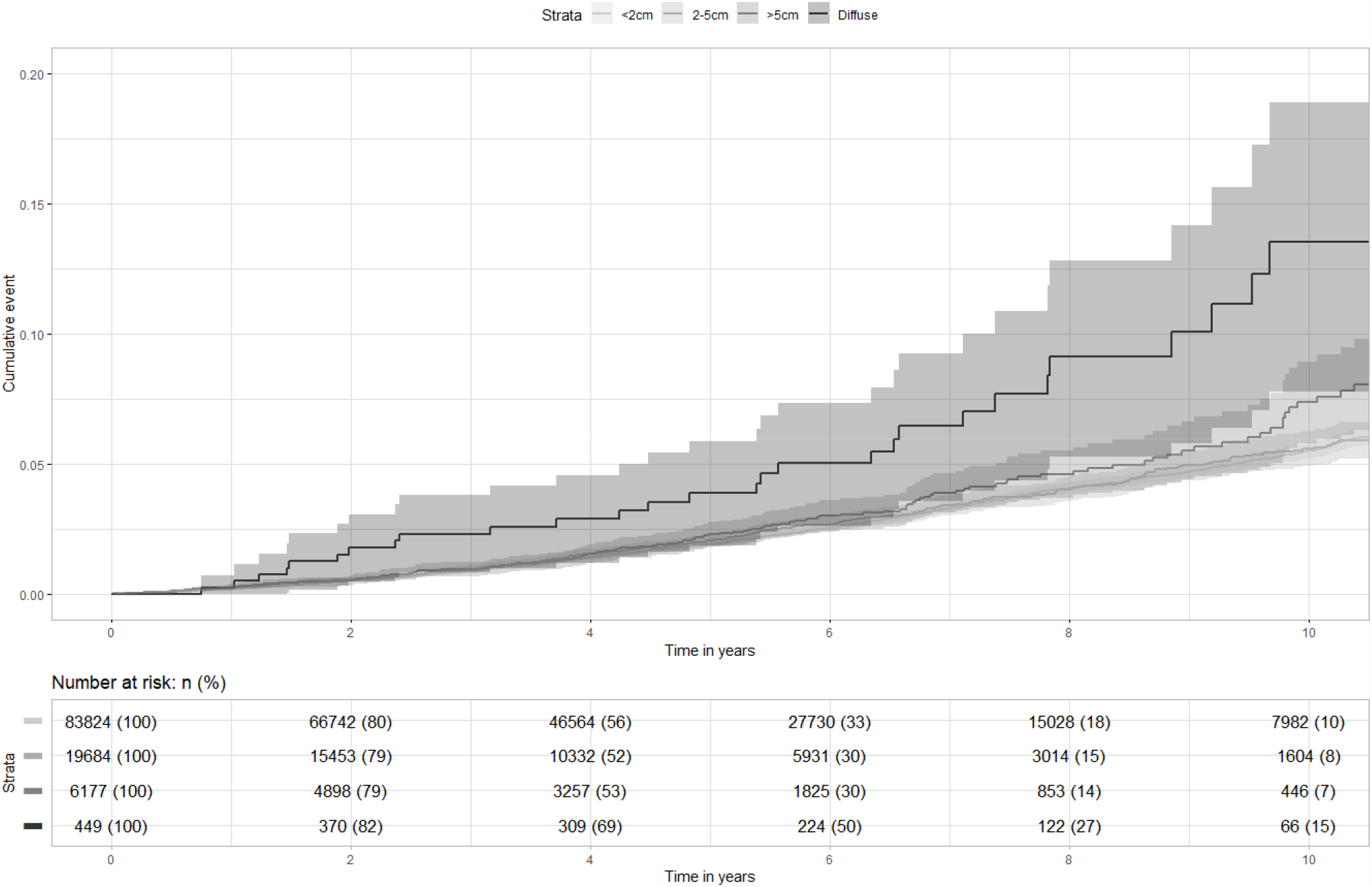
Cumulative all-cause mortality among DCIS patients in NCDB categorized by lesion size including diffuse type disease.

Among patients with diffuse disease, 37/188 (19.7%) underwent lumpectomy, as compared to 46918/70093 (66.9%) of patients without diffuse disease. On subgroup analysis of patients undergoing mastectomy for diffuse disease, the 10 year cumulative incidence of ACM for patients undergoing unilateral mastectomy was 15.0% and for those undergoing bilateral mastectomy was 2.5%, though this difference was not statistically significant on log rank test (p=0.11).

In univariate Cox regressions for ACM, survival was improved by administration of RT (HR=0.9, p=0.03), ET (HR=0.83, p=0.0003), age ≤50 (HR=0.41, p<0.0001), and “other” race (HR=0.52, p<0.0001) and was worsened by ER negative disease (HR=1.31, p<0.0001), lesion size 2 – 5 cm (HR=1.16, p=0.02), and black race (HR=1.87, p<0.0001). The only covariate found to violate proportional hazards assumption was administration of ET. In the multivariate Cox regression, overall survival was improved by mastectomy (HR=0.81, p=0.01), RT (HR=0.79, p=0.001), ET during the first 50 months after diagnosis (HR=0.71, p<0.0001), age ≤50 (HR=0.41, p<0.0001), and “other” race (HR=0.55, p=0.03). In contrast, overall survival was reduced by ER negative disease (HR=1.19, p=0.01), black race (HR=1.86, p<0.0001), lesion size 2 – 5 cm (HR=1.19, p=0.01) and diffuse lesion size (HR=2.00, p=0.03) (Table 3).

**Table 3.** Time varying univariate and multivariate Cox regression of NCDB patients for all-cause mortality.

|  | Univariate |  |  | Multivariate |  |  |
| --- | --- | --- | --- | --- | --- | --- |
|  | Hazard Ratio | 95% CI | p | Hazard Ratio | 95% CI | p |
| Surgery |  |  |  |  |  |  |
| Lumpectomy | Ref | - | - | Ref | - | - |
| Mastectomy | 0.96 | 0.86-1.07 | 0.45 | 0.81 | 0.69-0.95 | 0.01 |
| Radiation |  |  |  |  |  |  |
| Not administered | Ref | - | - | Ref | - | - |
| Administered | 0.90 | 0.81-0.99 | 0.03 | 0.79 | 0.68-0.91 | 0.001 |
| Endocrine therapy<sup>†</sup> |  |  |  |  |  |  |
| Not administered | Ref | - | - | Ref | - | - |
| Administered (≤50 months) | 0.69 | 0.59-0.80 | <0.0001 | 0.71 | 0.60-0.83 | <0.0001 |
| Administered (>50 months) | 0.98 | 0.85-1.13 | 0.79 | 1.04 | 0.90-1.22 | 0.57 |
| Estrogen receptor |  |  |  |  |  |  |
| Positive or borderline | Ref | - | - | Ref | - | - |
| Negative | 1.31 | 1.16-1.49 | <0.0001 | 1.19 | 1.04-1.38 | 0.01 |
| Race |  |  |  |  |  |  |
| White | Ref | - | - | Ref | - | - |
| Black | 1.87 | 1.64-2.12 | <0.0001 | 1.86 | 1.64-2.12 | <0.0001 |
| Other | 0.52 | 0.39-0.69 | <0.0001 | 2.00 | 0.41-0.73 | 0.03 |
| Age |  |  |  |  |  |  |
| >50 | Ref | - | - | Ref | - | - |
| ≤50 | 0.41 | 0.37-0.45 | <0.0001 | 0.41 | 0.37-0.46 | <0.0001 |
| Size |  |  |  |  |  |  |
| ≤2 cm | Ref | - | - | Ref | - | - |
| 2 – 5 cm | 1.16 | 1.02-1.32 | 0.02 | 1.19 | 1.03-1.35 | 0.01 |
| >5 cm | 1.12 | 0.91-1.40 | 0.28 | 1.15 | 0.92-1.43 | 0.02 |
| Diffuse | 1.85 | 0.99-3.45 | 0.05 | 2.00 | 1.07-3.75 | 0.03 |
| Grade |  |  |  |  |  |  |
| High | Ref | - | - | Ref | - | - |
| Intermediate | 1.02 | 0.91-1.14 | 0.74 | 1.08 | 0.96-1.21 | 0.21 |
| Low | 0.92 | 0.78-1.08 | 0.32 | 0.93 | 0.78-1.10 | 0.41 |

<sup>†</sup>Endocrine therapy was the only covariate violating the proportional hazards assumption and consequently was the only covariate for which a time-varying coefficient was utilized.

### SEER

A total of 31,752 patients met inclusion criteria for the final competing risks regression. 23,650 patients had lesion sizes ≤ 2 cm, 6,216 had lesions 2-5 cm, 1,830 patients had lesions > 5 cm, and 56 patients had lesion sizes classified as diffuse. Median follow up time was 5.6 years (interquartile range 3-8.75 years). Median age at diagnosis was 58.

Cumulative incidence of BCM at 10 years was 5.0% for diffuse lesions (3 diffuse patients total suffered BCM), 3.6% for patients with microinvasive disease, 2.3% for lesions >5 cm, 1.4% for lesions 2-5 cm and 1.5% for lesions <2 cm (Figure 3). When non-microinvasive invasive breast cancer cases of any size with no lymph node or metastatic involvement were included in the analysis, their 10 year BCM for comparison was 6.8%. Of note, of the three patients with diffuse type DCIS who suffered BCM, none had a documented second invasive breast event, similar to the patients observed in the UCSD cancer registry.

**Figure 3.**
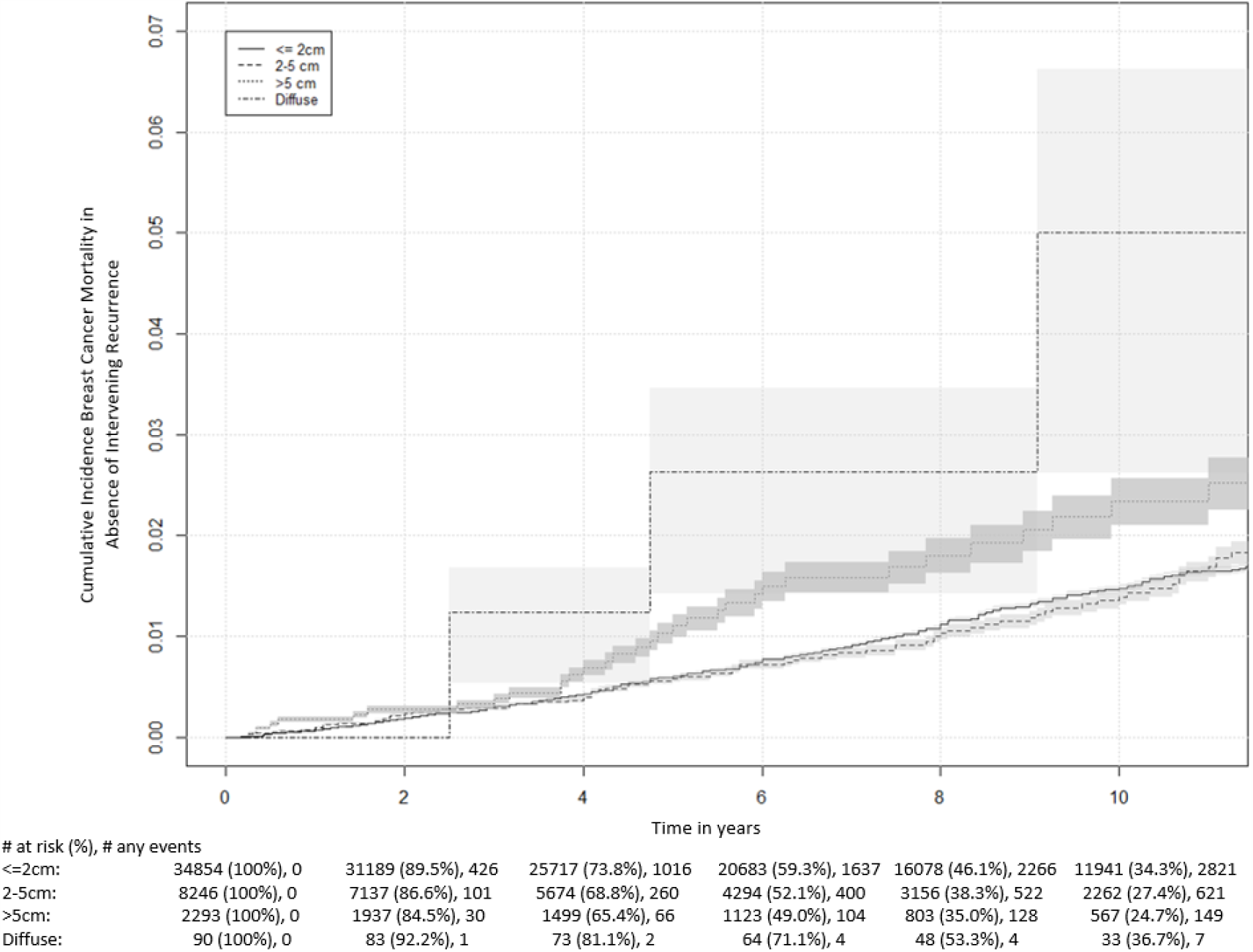
Cumulative incidence of breast cancer mortality among patients in SEER 18 with non-breast mortality as a competing event.

On competing risks regression, lower BCM was seen with adjuvant RT (HR=0.42, p<0.0001), mastectomy (HR=0.54, p=0.0006), and age ≤ 50 (HR=0.54, p=0.0004). In contrast, higher BCM was observed with ER negative disease (HR=1.65, p=0.001), black race (HR=2.58,p<0.0001), lesion size > 5 cm (HR=1.81, p=0.01) and diffuse disease (HR=4.33, p=0.04). On time-varying multivariate analysis, breast cancer specific survival was improved with administration of RT during the first 120 months after diagnosis (HR=0.36, p<0.0001), mastectomy (HR=0.47, p=0.0002), age ≤ 50 (HR=0.55, p=0.0007), and was worsened by black race during the first 45 months (HR=4.20, p<0.0001) and 45-110 months (HR=2.04, p=0.004) after diagnosis, as well as lesion size >5 cm(HR=1.76, p=0.03) and diffuse lesion distribution (HR=5.28, p=0.02) (Table 4).

**Table 4.** Time varying univariate and multivariate competing risks regression of SEER 18 patients for breast-cancer specific mortality with non-breast mortality treated as a competing event.

|  | Univariate |  |  | Multivariate |  |  |
| --- | --- | --- | --- | --- | --- | --- |
|  | HR | 95% CI | p | HR | 95% CI | p |
| Treatment |  |  |  |  |  |  |
| Lumpectomy | Ref | Ref | - | Ref | Ref | - |
| Lumpectomy + RT<br>(≤120 mo) | 0.52 | 0.39-0.69 | <0.0001 | 0.36 | 0.26-0.50 | <0.0001 |
| Lumpectomy + RT<br>(120-160 mo) | 2.33 | 0.75-7.27 | 0.14 | 1.65 | 0.53-5.19 | 0.39 |
| Lumpectomy + RT<br>(>160 mo) | 0.65 | 0.11-3.74 | 0.63 | 0.46 | 0.07-2.79 | 0.39 |
| Mastectomy | 0.98 | 0.72-1.32 | 0.87 | 0.47 | 0.32-0.70 | 0.0002 |
| Age |  |  |  |  |  |  |
| >50 | Ref | Ref | - | Ref | Ref | - |
| ≤50 | 0.54 | 0.38-0.76 | 0.0004 | 0.55 | 0.39-0.78 | 0.0007 |
| Size |  |  |  |  |  |  |
| ≤2 cm | Ref | Ref | - | Ref | Ref | - |
| 2 – 5 cm | 0.90 | 0.63-1.28 | 0.55 | 0.90 | 0.62-1.32 | 0.59 |
| >5 cm | 1.87 | 1.19-2.95 | 0.01 | 1.76 | 1.07-2.89 | 0.03 |
| Diffuse | 5.21 | 1.30-20.85 | 0.02 | 5.28 | 1.25-22.22 | 0.02 |
| Race |  |  |  |  |  |  |
| White | Ref | Ref | - | Ref | Ref | - |
| Black (≤45 mo) | 4.17 | 2.64-6.58 | <0.0001 | 4.20 | 2.66-6.63 | <0.0001 |
| Black (45-110 mo) | 1.97 | 1.21-3.20 | 0.006 | 2.04 | 1.26-3.29 | 0.004 |
| Black (>110 mo) | 0.51 | 0.58-4.74 | 0.34 | 1.60 | 0.55-4.64 | 0.38 |
| Other | 0.64 | 0.40-1.01 | 0.06 | 0.80 | 0.50-1.28 | 0.36 |
| ER status |  |  |  |  |  |  |
| ER Positive | Ref | Ref | - | Ref | Ref | - |
| ER Negative (≤60<br>mo) | 0.70 | 1.33-3.05 | 0.001 | 2.05 | 1.32-3.19 | 0.002 |
| ER Negative (60-140 mo) | 0.20 | 0.74-2.03 | 0.43 | 1.31 | 0.79-2.19 | 0.29 |
| ER Negative (>140<br>mo) | 0.78 | 0.65-7.26 | 0.21 | 0.89 | 0.66-8.03 | 0.19 |
| Grade |  |  |  |  |  |  |
| High grade | Ref | Ref | - | Ref | Ref | - |
| Intermediate grade<br>(≤50 months) | 0.89 | 0.59-1.35 | 0.59 | 0.89 | 0.57-1.37 | 0.59 |
| Intermediate grade<br>(50-75 months) | 1.58 | 0.84-2.96 | 0.15 | 1.42 | 0.74-2.72 | 0.29 |
| Intermediate grade<br>(>75 months) | 1.48 | 0.96-2.29 | 0.08 | 1.33 | 0.85-2.07 | 0.21 |
| Low grade | 0.57 | 0.35-0.94 | 0.03 | 0.59 | 0.35-0.99 | 0.05 |

<sup>†</sup>Time-varying coefficients in the form of step functions were introduced for covariates violating the proportional hazards assumption, including lumpectomy with radiation therapy treatment group, black race, ER negative disease, and intermediate tumor grade.

## Discussion

Here we demonstrate that diffuse GD DCIS is associated with a worse prognosis relative to non-diffuse GD DCIS. Importantly, by worse prognosis we specifically refer to development of DMD, BCM, and OS. This is in contrast to previous DCIS outcomes studies which commonly use in situ and invasive recurrence as the endpoints of interest [31-34]. The emphasis on recurrence in most studies is likely due to the low BCM rate among DCIS patients.

While the numbers of patients with diffuse GD type DCIS in SEER and NCDB were very small, this is almost certainly related to inconsistent documentation regarding diffuse GD. For example, in the cancer registry data at UCSD, none of the 20 patients whom we were able to classify as diffuse were coded as such for submission to national cancer registries. Given that 20 of the 113 patients with classifiable growth distribution who underwent mastectomy had diffuse GD, 27-56% of DCIS patients undergo mastectomy for their disease, and 25.5% of patients in our SEER analysis with diffuse GD underwent lumpectomy rather than mastectomy, the proportion of DCIS patients with diffuse GD is almost certainly much higher. In Tot’s series of DCIS cases, for example, 36/108 (33.3%) of lesions were diffuse [35].

Furthermore, it is important to note that all of the patients in both our cancer registry analysis and in SEER developed DMD or suffered BCM in the absence of an intervening invasive event. This makes it highly likely that our estimated rate of BCM in SEER is an underestimate, as SEER instructs tumor registrars to recode initial entries for patients who develop DMD after pure DCIS as invasive type lesions, which reduces the apparent BCM in the absence of intervening invasive events without affecting the BCM rate after an invasive intervening recurrence [36].

While our outcomes analysis cannot provide a mechanism for the phenomenon by which these patients develop DMD, even years after bilateral mastectomies, we consider several possibilities. The simplest is that given the association in our cancer registry analysis of diffuse GD with large extent of disease, it may be more likely that a focus of invasion is missed, either spatially by sectioning or review of histopathology, or temporally with respect to timing of surgery. An alternative explanation is that the scattered microscopic foci of disease in diffuse GD may represent distinct transformation events leading to increased phenotypic heterogeneity. This would increase the chances not only of the development of a focus or foci of invasion, but also a greater number of cells with invasive potential that would increase the chances of at least one such cell lines having the potential to adapt to a distant metastatic microenvironmental niche [37, 38].

Given the retrospective nature of our study, association cannot be presumed to be causative, but it is worth noting that unilateral mastectomy was associated with worse outcomes for diffuse patients in all three studies: 2/3 patients who developed diffuse GD in our cancer registry underwent unilateral mastectomy (the third had a bilateral mastectomy), 2/3 patients who died of breast cancer in SEER after diffuse DCIS had unilateral mastectomy (the remaining third had a unilateral lumpectomy), and the 10-year cumulative overall mortality in NCDB for diffuse patients was 15% among patients undergoing unilateral mastectomy vs. 2.5% among patients undergoing bilateral mastectomy. While causation cannot be demonstrated, we believe that given the very poor outcomes, it would be reasonable to recommend for patients with diffuse type disease already requiring unilateral mastectomy a bilateral mastectomy instead. Importantly, we think that systemic therapy is likely more important for these patients, but quality prospective data would be required before recommending such treatment for basal-like DCIS patients, given the significant risks associated with chemotherapy.

Overall strengths of the study included the relevant findings specific to each study (DMD in cancer registry, OS in NCDB, and BCM in SEER) and the utilization of time-varying coefficients in the form of step functions in all three analyses and the utilization of competing risks in the SEER analysis, which, unlike time-varying covariates [39], retain their relationship to the cumulative incidence functions in a given time frame. Overall weaknesses included the lack of central pathology review, reliance on SEER and NCDB for classification of diffuse type disease, and the retrospective nature of the study preventing such options as randomization and standardized patient follow up and post-operative screening. SEER and NCDB guidelines both involve pathology reports as the priority for size estimates, and we filtered our results for only patients undergoing surgery with known lesion sizes. However, in the national databases unlike in our local cancer registry, we are unable to confirm that patients classified as having diffuse disease are consistently referring to the same histopathologic features.

Regarding the subsections of our analyses, different portions had different strengths and weaknesses. Our analysis of mastectomy patients in the UCSD cancer registry had the highest quality data since we chart reviewed all variables, allowed for characterization of growth distribution by pathology reports regarding subgross pathology, and had the longest median follow up time; this data did, however, have the smallest sample size. SEER had a much larger sample size and allowed for analysis of several outcomes of interest including overall and breast cancer specific survival, but did not have data regarding endocrine therapy. NCDB had the largest sample size but only offered the outcome of OS.

In conclusion, diffuse GD represents a subtype of DCIS with very poor prognosis with respect to metastasis and survival. Among patients requiring mastectomy, bilateral rather than unilateral may be advisable. Further work is needed to elucidate the best treatment course for these patients.

## Data Availability

SEER and NCDB data are maintained by NIH and American College of Surgeons, respectively. Data in the single institution analysis are not publicly available as they contain PHI but the records are obtainable for research parties with IRB approval through the UCSD HRPP.

## Acknowledgements

We acknowledge the National Cancer Institute, specifically the SEER team, for granting us access to SEER data. We acknowledge the American College of Surgeons, specifically the NCDB team, for granting us access to the NCDB breast data.

## References

1. Punglia, R.S., et al., Epidemiology, Biology, Treatment, and Prevention of Ductal Carcinoma In Situ (DCIS). JNCI Cancer Spectr, 2018. 2(4): p. pky063.

2. Early Breast Cancer Trialists’ Collaborative, G., et al., Overview of the randomized trials of radiotherapy in ductal carcinoma in situ of the breast. J Natl Cancer Inst Monogr, 2010. 2010(41): p. 162–77.

3. Barrio, A.V. and K.J. Van Zee, Controversies in the Treatment of Ductal Carcinoma in Situ. Annu Rev Med, 2017. 68: p. 197–211.

4. Fisher, B., et al., Lumpectomy and radiation therapy for the treatment of intraductal breast cancer: findings from National Surgical Adjuvant Breast and Bowel Project B-17. J Clin Oncol, 1998. 16(2): p. 441–52.

5. Donker, M., et al., Breast-conserving treatment with or without radiotherapy in ductal carcinoma In Situ: 15-year recurrence rates and outcome after a recurrence, from the EORTC 10853 randomized phase III trial. J Clin Oncol, 2013. 31(32): p. 4054–9.

6. Holmberg, L., et al., Absolute risk reductions for local recurrence after postoperative radiotherapy after sector resection for ductal carcinoma in situ of the breast. J Clin Oncol, 2008. 26(8): p. 1247–52.

7. Cuzick, J., et al., Effect of tamoxifen and radiotherapy in women with locally excised ductal carcinoma in situ: long-term results from the UK/ANZ DCIS trial. Lancet Oncol, 2011. 12(1): p. 21–9.

8. McGuire, K.P., et al., Are mastectomies on the rise? A 13-year trend analysis of the selection of mastectomy versus breast conservation therapy in 5865 patients. Ann Surg Oncol, 2009. 16(10): p. 2682–90.

9. Silverstein, M.J., et al., Ten-year results comparing mastectomy to excision and radiation therapy for ductal carcinoma in situ of the breast. Eur J Cancer, 1995. 31A(9): p. 1425–7.

10. Ward, E.M., et al., Cancer statistics: Breast cancer in situ. CA Cancer J Clin, 2015. 65(6): p. 481–95.

11. Narod, S.A., et al., Breast Cancer Mortality After a Diagnosis of Ductal Carcinoma In Situ. JAMA Oncol, 2015. 1(7): p. 888–96.

12. Hwang, E.S., et al., The COMET (Comparison of Operative versus Monitoring and Endocrine Therapy) trial: a phase III randomised controlled clinical trial for low-risk ductal carcinoma in situ (DCIS). BMJ Open, 2019. 9(3): p. e026797.

13. Francis, A., et al., Addressing overtreatment of screen detected DCIS; the LORIS trial. Eur J Cancer, 2015. 51(16): p. 2296–303.

14. Elshof, L.E., et al., Feasibility of a prospective, randomised, open-label, international multicentre, phase III, non-inferiority trial to assess the safety of active surveillance for low risk ductal carcinoma in situ - The LORD study. Eur J Cancer, 2015. 51(12): p. 1497–510.

15. Wadsten, C., et al., Risk of death from breast cancer after treatment for ductal carcinoma in situ. Br J Surg, 2017. 104(11): p. 1506–1513.

16. Narod, S.A. and V. Sopik, Is invasion a necessary step for metastases in breast cancer? Breast Cancer Res Treat, 2018. 169(1): p. 9–23.

17. Hollingsworth, A.B., Is invasion a necessary step for metastases in breast cancer? Narod SA, Sopik V. Breast Cancer Res Treat, 2018. 169(3): p. 633–637.

18. Osako, T., et al., Incidence and possible pathogenesis of sentinel node micrometastases in ductal carcinoma in situ of the breast detected using molecular whole lymph node assay. Br J Cancer, 2012. 106(10): p. 1675–81.

19. Lara, J.F., et al., The relevance of occult axillary micrometastasis in ductal carcinoma in situ: a clinicopathologic study with long-term follow-up. Cancer, 2003. 98(10): p. 2105–13.

20. Sanger, N., et al., Molecular Markers as Prognostic Factors in DCIS and Small Invasive Breast Cancers. Geburtshilfe Frauenheilkd, 2014. 74(11): p. 1016–1022.

21. Franken, B., et al., Circulating tumor cells, disease recurrence and survival in newly diagnosed breast cancer. Breast Cancer Res, 2012. 14(5): p. R133.

22. Gruber, I.V., et al., Relationship Between Hematogenous Tumor Cell Dissemination and Cellular Immunity in DCIS Patients. Anticancer Res, 2016. 36(5): p. 2345–51.

23. Silverstein, M.J., et al., A prognostic index for ductal carcinoma in situ of the breast. Cancer, 1996. 77(11): p. 2267–74.

24. Cheng, L., et al., Relationship between the size and margin status of ductal carcinoma in situ of the breast and residual disease. J Natl Cancer Inst, 1997. 89(18): p. 1356–60.

25. Neri, A., et al., ”Clinical significance of multifocal and multicentric breast cancers and choice of surgical treatment: a retrospective study on a series of 1158 cases”. BMC Surg, 2015. 15: p. 1.

26. Bonnier, P., et al., Prognostic factors in ductal carcinoma in situ of the breast: results of a retrospective study of 575 cases. The Association for Research in Oncologic Gynecology. Eur J Obstet Gynecol Reprod Biol, 1999. 84(1): p. 27–35.

27. Tot, T., Clinical relevance of the distribution of the lesions in 500 consecutive breast cancer cases documented in large-format histologic sections. Cancer, 2007. 110(11): p. 2551–60.

28. Tot, T., et al., Breast cancer multifocality, disease extent, and survival. Hum Pathol, 2011. 42(11): p. 1761–9.

29. Tot, T., et al., Molecular phenotypes of unifocal, multifocal, and diffuse invasive breast carcinomas. Patholog Res Int, 2010. 2011: p. 480960.

30. Fine, J.P. and R.J. Gray, A Proportional Hazards Model for the Subdistribution of a Competing Risk. Journal of the American Statistical Association, 1999. 94(446): p. 496–509.

31. van de Vijver, M.J., Biological variables and prognosis of DCIS. Breast, 2005. 14(6): p. 509–19.

32. Silverstein, M.J., et al., Prognostic classification of breast ductal carcinoma-in-situ. Lancet, 1995. 345(8958): p. 1154–7.

33. Nofech-Mozes, S., et al., Prognostic and predictive molecular markers in DCIS: a review. Adv Anat Pathol, 2005. 12(5): p. 256–64.

34. Pinder, S.E., Ductal carcinoma in situ (DCIS): pathological features, differential diagnosis, prognostic factors and specimen evaluation. Mod Pathol, 2010. 23 Suppl 2: p. S8–13.

35. Tot, T., DCIS, cytokeratins, and the theory of the sick lobe. Virchows Arch, 2005. 447(1): p. 1–8.

36. O’Keefe, T.J. and A.M. Wallace, Surveillance, Epidemiology, and End Results program underestimates breast cancer-specific mortality after ductal carcinoma in situ diagnosis. Breast Cancer Res Treat, 2020.

37. Hosseini, H., et al., Early dissemination seeds metastasis in breast cancer. Nature, 2016. 540(7634): p. 552–558.

38. Asioli, S., et al., The impact of field cancerization on the extent of duct carcinoma in situ (DCIS) in breast tissue after conservative excision. Eur J Surg Oncol, 2016. 42(12): p. 1806–1813.

39. Austin, P.C., A. Latouche, and J.P. Fine, A review of the use of time-varying covariates in the Fine-Gray subdistribution hazard competing risk regression model. Stat Med, 2020. 39(2): p. 103-113. size including diffuse type disease.

